# Longitudinal course of inflammatory-cognitive subgroups across first treatment severe mental illness and healthy controls

**DOI:** 10.1101/2024.04.08.24305500

**Authors:** Linn Sofie Sæther, Thor Ueland, Beathe Haatveit, Anja Vaskinn, Camilla Bärthel Flaaten, Christine Mohn, Monica B. E.G. Ormerod, Pål Aukrust, Ingrid Melle, Nils Eiel Steen, Ole A. Andreassen, Torill Ueland

## Abstract

**Background:** While inflammation is associated with cognitive impairment in severe mental illnesses (SMI), there is substantial heterogeneity and evidence of transdiagnostic subgroups across schizophrenia (SZ) and bipolar (BD) spectrum disorders. There is however, limited knowledge about the longitudinal course of this relationship.

**Methods:** Systemic inflammation (C-Reactive Protein, CRP) and cognition (nine cognitive domains) was measured from baseline to 1 year follow-up in first treatment SZ and BD (n=221), and healthy controls (HC, n=220). Linear mixed models were used to evaluate longitudinal changes separately in CRP and cognitive domains specific to diagnostic status (SZ, BD, HC). Hierarchical clustering was applied on the entire sample to investigate the longitudinal course of transdiagnostic inflammatory-cognitive subgroups.

**Results:** There were no case-control differences or change in CRP from baseline to follow-up. We confirm previous observations of case-control differences in cognition at both time-points and domain specific stability/improvement over time regardless of diagnostic status. We identified transdiagnostic inflammatory-cognitive subgroups at baseline with differing demographics and clinical severity. Despite improvement in cognition, symptoms and functioning, the higher inflammation – lower cognition subgroup (75% SZ; 48% BD; 38% HC) had sustained inflammation and lower cognition, more symptoms, and lower functioning (SMI only) at follow-up. This was in comparison to a lower inflammation – higher cognition subgroup (25% SZ, 52% BD, 62% HC), where SMI participants showed cognitive functioning at HC level with a positive clinical course.

**Conclusions:** Our findings support heterogenous and transdiagnostic inflammatory-cognitive subgroups that are stable over time, and may benefit from targeted interventions.

## Introduction

Cognitive impairment is a central feature of severe mental illnesses (SMI), such as schizophrenia (SZ) and bipolar (BD) spectrum disorders (McCleery and Nuechterlein, 2019; Stainton et al., 2023). While highly prevalent, there is considerable heterogeneity in cognitive symptoms, ranging from mild to severe (Catalan et al., 2024; Haatveit et al., 2023; Lee et al., 2024; Van Rheenen et al., 2017; Wenzel et al., 2023). Numerous studies have identified transdiagnostic cognitive subgroups that are associated with different neurobiological characteristics, as well as clinical- and functional outcomes (Bora et al., 2023; Cowman et al., 2021; Lewandowski, 2020; Vaskinn et al., 2020; Wenzel et al., 2021). For instance, cognitive subgroups with severe impairment typically have more symptoms and lower functioning (Miskowiak et al., 2023; Vaskinn et al., 2020), brain abnormalities as assessed by magnetic resonance imaging (de Zwarte et al., 2020; Wenzel et al., 2023, 2021; Wolfers et al., 2018; Woodward and Heckers, 2015), and higher levels of systemic inflammation (Pan et al., 2020, Watson et al., 2023). Evidence further suggests that cognitive functioning remains relatively stable throughout the illness course in both SZ and BD (Bora and Özerdem, 2017; Catalan et al., 2024; Ehrlich et al., 2022; Flaaten et al., 2022, 2023a, 2023b). Developing successful personalized treatments is contingent on increasing our understanding of the causes and maintenance of cognitive impairment in SMI.

Current pharmacotherapies targeting symptom relief in SMI have limited effects on cognition, which may have a different underlying pathophysiology (Howes et al., 2024; McCutcheon et al., 2023). Evidence suggests immune- and inflammatory-related abnormalities, which are well documented across the psychosis spectrum (Andreassen et al., 2023; Benros et al., 2014; Goldsmith et al., 2016; Steen et al., 2023; Webster, 2023), are associated with cognitive impairment (Jovasevic et al., 2024; Morozova et al., 2022; Rosenblat et al., 2015; Wang et al., 2022). Dysregulated systemic levels of inflammatory markers have been observed in first-episode and chronic stages of SMI (Halstead et al., 2023; Perry et al., 2021), including in medication naïve patients (Dunleavy et al., 2022; Fernandes et al., 2016; van den Ameele et al., 2016). The most extensively studied and reliable marker of systemic inflammation in SMI is C-Reactive Protein (CRP), in part due its low-cost and global accessibility at routine medical laboratories (Clyne and Olshaker, 1999; Ullah et al., 2021). CRP levels fluctuate in response to change in inflammatory status and may be used to infer whether low-grade systemic inflammation is associated with cognitive impairment. In fact, increased levels of CRP have been consistently reported in SZ and BD relative to healthy controls, and previously found to be modestly associated with clinical- and cognitive characteristics (Fernandes et al., 2016; Fond et al., 2018; Halstead et al., 2023; Jacomb et al., 2018; Johnsen et al., 2016; Lestra et al., 2022; Millett et al., 2021; Misiak et al., 2018; Patlola et al., 2023).

It is increasingly clear that only a subset of individuals with SMI show signs of increased systemic inflammation (Bishop et al., 2022; Chen et al., 2024; Miller and Goldsmith, 2019), partly explaining mixed or weak associations between inflammatory markers and cognition in case-control studies (Bora, 2019; Miller and Goldsmith, 2019; Morrens et al., 2022). This is also in line with genetic findings of mixed effect directions, which includes higher load of increasing and decreasing genetic variants for CRP in SMI (Hindley et al., 2023). Similar to findings on cognitive subgroups (Bora et al., 2023; Cowman et al., 2021; Lewandowski, 2020; Wenzel et al., 2023, 2021), the higher-inflammation subtype is associated with more adverse neurobiological and clinical outcomes, and is associated with lower cognitive functioning (Boerrigter et al., 2017; Fillman et al., 2016; Lizano et al., 2023, 2020; Millett et al., 2021; Nettis et al., 2019; Zhang et al., 2022). A common observation is that a larger proportion of individuals with SMI compared to control participants, belong to a higher-inflammation subtype (Boerrigter et al., 2017; Fillman et al., 2016; Lizano et al., 2020). Including both SMI and control participants when using unsupervised clustering techniques allows for evaluation of similarities and differences across phenotypes, regardless of diagnostic status.

Recent evidence from machine learning suggests higher accuracy of case-control prediction when both cognition and inflammatory markers are evaluated together (Fernandes et al., 2020). Using hierarchical clustering, we recently identified a transdiagnostic subgroup with cognitive impairment and higher inflammation using different immune and inflammatory marker panels (Sæther et al., 2023, 2024). This subgroup also had more symptoms and lower functioning, compared to a subgroup with milder impairments and lower inflammation. The clinical relevance of these subgroups remains to be determined, and longitudinal studies are essential to address if these subgroups are trait or state phenomenon. Longitudinal studies on subgroups based on cognition suggest stability over time for both SZ and BD (Ehrlich et al., 2022; Flaaten et al. 2022; Lim et al., 2021). Longitudinal studies of inflammatory markers, including CRP, are in general scarce, and most of them focus on the effects of antipsychotic treatment in SZ cohorts only (Fathian et al., 2022; Feng et al., 2020; Meyer et al., 2009). Evidence based on a few studies suggests a diminished correlation between CRP and cognition after 6 weeks of admittance to hospital with acute psychosis (Johnsen et al., 2016), and an early drop in CRP level may predict improved cognitive functioning after 6 months (Fathian et al., 2019). To our knowledge, no previous study has evaluated temporal characteristics subgroups based on both inflammation and cognition in SMI and controls.

The current study aimed to elucidate the longitudinal course of systemic inflammation and cognition in first treatment SMI (SZ=133, BD=88), and healthy controls (n=220). Inflammation was assessed with CRP, and cognition with nine core domains including fine-motor speed, psychomotor processing speed, mental processing speed, attention, verbal learning, verbal memory, semantic fluency, working memory and cognitive control, at baseline and ∼12 months later (first year of adequate treatment). We first investigated the specific trajectories of CRP levels and cognitive domains associated with diagnostic status (SZ, BD, HC), using separate linear mixed models. Based on our findings from previous overlapping samples, we expect domain-specific stability or improvement over the first year of treatment in SMI and HC (Demmo et al., 2017; Engen et al., 2019; Haatveit et al., 2015). The trajectory of CRP levels from baseline to 1 year follow-up in first treatment SZ and BD is, however, unknown. Based on a similar approach to our previous work (Sæther et al., 2024, 2023), we used hierarchical clustering to identify transdiagnostic inflammatory-cognitive subgroups using CRP and a cognitive composite score at baseline. The subgroups were assessed longitudinally across demographic, clinical, and cognitive measures.

## Methods

### Sample

This study is part of the ongoing Thematically Organized Psychosis (TOP)-study. Participants meeting the Diagnostic Manual of Mental Disorders (DSM)-Ⅳ criteria for schizophrenia or bipolar spectrum disorders are continuously recruited from in- and out-patient psychiatric units in the larger Oslo area. Healthy controls (HC) from the same catchment area are randomly chosen using statistical records and invited by letter. Exclusion criteria for all participants are: 1) age <18 or >65, 2) moderate/severe head injury, 3) severe somatic/neurological disorder, 4) not fluent in a Scandinavian language, 5) IQ<70. HC are excluded in the case of drug dependency, history of mental illness, or relatives with SMI. Any participant (SMI and HC) with signs of acute infection at baseline and/or follow-up (CRP>10 mg/L) was excluded. This study included SMI participants who at baseline was within the first 12 months of starting their first adequate treatment of SZ or BD spectrum disorder. Participants had to have follow-up assessment 6 months to 1.5 year later (mean=400 days), with relatively complete cognitive assessment at both time points, and blood samples taken at both time points. Baseline assessments were conducted between 2004-2020, and follow-up assessments between 2005-2021. The final sample included n=133 SZ spectrum (schizophrenia=76, schizophreniform=13, schizoaffective=8, psychosis not otherwise specified=36), n=88 BD spectrum (bipolar I=53, bipolar II= 30, bipolar not otherwise specified=5) and n=220 healthy controls. Due to selection criteria the retention rate for this study was not possible to determine. However, the retention rate for one-year follow-up of cognitive assessment in the TOP-study has previously been reported to be 53-66%, with little or no difference in clinical or demographic characteristics between those eligible for follow-up versus completers (Demmo et al., 2017; Engen et al., 2019). All participants provided informed consent and the study was approved by the Regional Ethics Committee.

### Clinical assessments

The Structured Clinical Interview for DSM-Ⅳ axis 1 disorders (SCID-Ⅰ)(First et al., 1995) was administered by trained clinical psychologists or physicians. The Positive and Negative Syndrome Scale (PANSS) was used to assess symptoms according to the five-factor model including positive, negative, disorganized/concrete, excited and depressed symptoms (Kay et al., 1987; Wallwork et al., 2012). Manic symptoms were assessed with the Young Mania Rating Scale (YMRS) (Young et al., 1978). Level of functioning was assessed with the split version of the Global Assessment of Functioning scale (GAF F, GAF S; Pedersen et al., 2007). Duration of untreated psychosis (DUP) was estimated as time of onset from psychotic symptoms until start of first adequate treatment. The average time between physical examination (blood sampling, height/weight), and cognitive assessment was 4.2 days for baseline and 5.3 days at follow-up. The defined daily dose (DDD) of psychopharmacological treatment (antipsychotics, antidepressants, antiepileptics and lithium) was determined according to World Health Organization guidelines (https://www.whocc.no/atc_ddd_index). Somatic medication use (yes/no) in the SMI group is provided in Table S1.

### Cognitive assessments

Trained clinical psychologists or research personnel administered one of two test batteries: Battery 1 (from 2004-2012) or Battery 2 (from 2012). The test batteries included different tests of equivalent cognitive functions, as well as some identical measures. Thus, to ensure the highest possible N, corresponding tests from the two batteries were standardized separately (Z-scores) before combining to cover nine cognitive domains: *Fine-motor speed, psychomotor processing speed, mental processing speed, attention, verbal learning, verbal memory, semantic fluency, working memory and cognitive control*. We have previously shown robust between-battery correspondence of test performance for SZ, BD and HC (Sæther et al., 2024). The cognitive batteries consisted of tests from the MATRICS Consensus Cognitive Battery (MCCB) (Nuechterlein et al., 2008), Halstead-Reitan (Klove, 1963), the Wechsler Adult Intelligence Scale (WAIS-III) (Wechsler, 1997), Delis Kaplan Executive Functioning System (D-KEFS) (Delis et al., 2001), the California Verbal Learning Test (CVLT-II) (Delis et al., 1987), and the Hopkins Verbal Learning Test-Revised (HVLT-R) (Benedict et al., 1998). We assessed intellectual functioning with the Matrix Reasoning and Vocabulary subtests from the Wechsler Abbreviated Scale of Intelligence (WASI) (Wechsler, 2011). See Table S2 for an overview of tests.

### Blood sampling

Blood was sampled from the antecubital vein in EDTA vials and stored at 4°C overnight before transport to the hospital central laboratory the next day. The samples (2x9 ml EDTA tubes) were centrifuged at 1800 g for 15 minutes, and isolated plasma was stored at -80 °C in multiple aliquots. Blood samples were analysed for CRP by a particle enhanced immunoturbidimetric method with a Cobas 8000 instrument (Roche Diagnostics, Basel Switzerland) at the Department of Medical Biochemistry, Oslo University Hospital, Oslo, Norway.

### Statistical procedure

#### Data preprocessing, sample, and clinical characteristics

Data preprocessing, statistical analyses and visualization of results were conducted in the R-environment (https://www.r-project.org/; v.4.2.0, main R-packages reported in Supplementary Methods 1). Cognitive data was standardized (Z-scores) based on the HC group mean and standard deviation (SD) at baseline, and CRP was log-10 transformed. A cognitive composite score was computed as the mean score across cognitive domains for participants with baseline data in at least 5 cognitive domains. Sample and clinical characteristics were compared across groups using Kruskal-Wallis rank sum test and pairwise permutation (*n*=10000) based t-tests for continuous variables, and chi-squared tests for categorical variables. All analyses were adjusted for multiple comparisons using Bonferroni correction. An overview of the number of observations for CRP and all cognitive domains at baseline and follow-up, as well as descriptive statistics for these can be found in Table S3-S4. Correlations between CRP and cognitive domains at baseline and follow-up are found in Fig. S1.

#### Linear Mixed Models

Linear mixed models were used to analyze group-level changes over time separately for CRP and each cognitive domain in order to account for individual variability and repeated measures within subjects. We included sex and age as covariates as they may impact cognition in the cognitive model, and sex, age, and BMI as covariates in the CRP model as they may influence CRP (in the CRP model). We used the following formula for cognitive data:

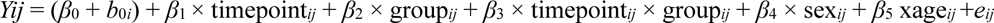

Where Y_ij_ is the cognitive score for participant i=1…441 at time j=0…1, *β* signifies fixed effects, b random effects (random intercept for each unique ID), and *e* the residual error term. The same model structure was used for CRP, with the addition of BMI as a covariate.

#### Hierarchical clustering

We used hierarchical clustering to identify subgroups based on inflammation and cognition in a subsample of participants with available CRP and a cognitive composite score at baseline (SZ=121, BD=87, HC=216). In brief, we 1) generated a Euclidian distance matrix, 2) evaluated the optimal linkage method based on the agglomerative coefficient (average, single, complete, Ward’s), 3) determined the optimal number of clusters by inspecting the average silhouette index, 4) tested the presence of clusters using a previously described data simulation procedure (Dinga et al., 2019), and 5) evaluated the stability of the cluster solution using a resampling procedure (bootstrapping). A Jaccard similarity index for clustering stability was computed with an index >0.7 (70%) was considered stable. We compared the subgroups on inflammation, cognition, sample (age, sex, education, IQ, BMI), clinical, and functional characteristics, at baseline and follow-up using Welch’s t-tests (effect sizes: Cohen’s *d*). In the case of sustained subgroup differences in any of the sample/clinical/functional characteristics, we investigated the effect of time, and potential subgroup differences in change over time (e.g. change scores, Λ1Y=Y_1_-Y_0_), using Wilcoxon signed-rank tests. All comparisons were corrected for multiple comparisons (Bonferroni).

### Code availability

Main analysis code/scripts are available at: https://osf.io/ek68q/

## Results

### Sample and clinical demographics

Sample and clinical characteristics at baseline are provided in Table 1. See Table S5 for clinical characteristics at follow-up.

**Table 1.** Sample and clinical characteristics at baseline.

| Characteristics | SZ<br>(N = 133)<sup>1</sup> | BD<br>(N = 88)<sup>1</sup> | HC<br>(N = 220)<sup>1</sup> | *p*-value<sup>2</sup> | Group<br>comparisons<sup>3</sup> |
| --- | --- | --- | --- | --- | --- |
| Age | 26.2 (6.8) | 28.6 (8.7) | 33.6 (9.3) | <0.001 | SZ, BD < HC |
| Sex (% female) | 52 (39) | 52 (59) | 107 (49) | 0.014 | SZ ~ BD, HC |
| Education (years) | 12.7 (2.8) | 13.6 (2.2) | 14.5 (2.1) | <0.001 | SZ, BD < HC |
| WASI IQ (2-subtests) | 102.9 (14.3) | 111.6 (11.9) | 113.8 (9.5) | <0.001 | SZ < BD, HC |
| BMI (kg/m<sup>2</sup>) | 24.2 (4.3) | 24.5 (4.3) | 24.8 (3.7) | ns | - |
| PANSS Negative | 12.9 (5.6) | 8.6 (3.1) |  | <0.001 | SZ > BD |
| PANSS Positive | 10.1 (4.0) | 5.7 (2.5) |  | <0.001 | SZ > BD |
| PANSS Disorganized | 5.5 (2.4) | 4.2 (1.7) |  | <0.001 | SZ > BD |
| PANSS Excited | 5.2 (1.6) | 5.0 (1.4) |  | ns | - |
| PANSS Depressed | 8.6 (2.9) | 8.1 (3.0) |  | ns | - |
| YMRS | 5.5 (4.9) | 3.2 (5.0) |  | <0.001 | SZ > BD |
| GAF Symptom | 43.2 (13.6) | 59.1 (10.7) |  | <0.001 | SZ < BD |
| GAF Function | 45.4 (14.1) | 55.9 (12.0) |  | <0.001 | SZ < BD |
| Days of untreated illness | 112.9 (193.9) | 59.2 (184.5) |  | <0.001 | SZ > BD |
| Antipsychotics, DDD | 1.3 (0.7) | 0.9 (0.7) |  | <0.001 | SZ > BD |
| Antidepressants, DDD | 1.4 (0.6) | 1.6 (0.9) |  | ns | - |
| Antiepileptics, DDD | 0.4 (0.2) | 0.8 (0.5) |  | 0.050 | SZ < BD |
| Lithium, DDD | - | 1.1 (0.3) |  | - | - |
| Total, DDD | 1.8 (1.1) | 1.7 (1.1) |  | ns | - |

<sup>1</sup>Mean (SD); n (%)<sup>2</sup>Kruskal-Wallis rank sum test; Pearson's Chi-squared test<sup>3</sup>Pairwise two-sample Permutation Test (for 3 groups)
Abbreviations: SZ, schizophrenia; BD, bipolar disorder; HC, healthy controls; WASI, Wechsler Abbreviated Scale of Intelligence; BMI, body mass index; PANSS, Positive and Negative Syndrome Scale; YMRS, Young Mania Rating Scale; GAF, Global Assessment of Functioning scale; DDD, defined daily dosage; ns, non-significant

### Inflammation and cognition over time comparing diagnostic status

As seen in Fig. 1, temporal assessment using linear mixed models suggested stable levels of CRP over time. There was no difference between SMI groups or HC at baseline or follow-up (Table 2), with a positive relationship between BMI and CRP (Table S6). For cognitive measures (Fig. S2), we confirm previous findings from studies using overlapping samples (Demmo et al., 2017; Engen et al., 2019; Flaaten et al., 2023b, 2023a, 2022; Haatveit et al., 2015), i.e. regardless of time-point, the cognitive scores remained attenuated in SMI, with SZ on average scoring ∼1 SD and BD ∼0.5 SD lower than HC. BD, however, had similar performance to HC on attention and semantic fluency at both time-points. Further, for all groups there was improvement in fine-motor speed, psychomotor speed, verbal learning, and cognitive control over time, whereas stability was observed for the remaining domains (mental speed, verbal memory, attention, semantic fluency, working memory). There was a significant time by group interaction for working memory, indicating improved performance over time for BD relative to HC. See Table S6 for extended model output.

**Figure 1.**
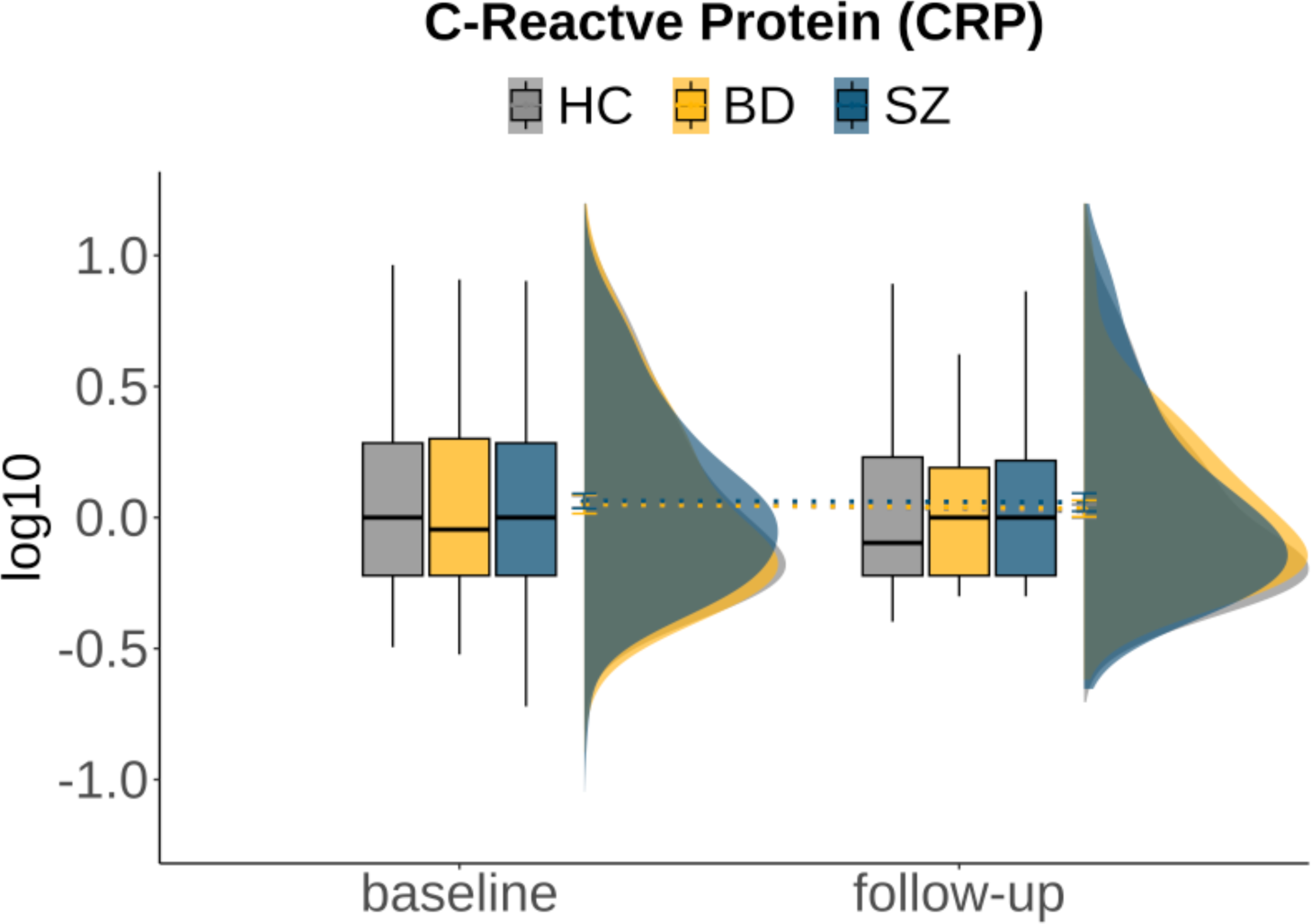
Inflammation (C-Reactive Protein, CRP) at baseline and follow-up between HC, BD and SZ. Boxplots (interquartile range separated by median line), density plot (kernel density estimate) and lines between mean scores (including error bars: ±SEM) shows no difference between HC, BD and SZ at baseline or follow-up and indicates stability of CRP-levels over time.

**Table 2.**
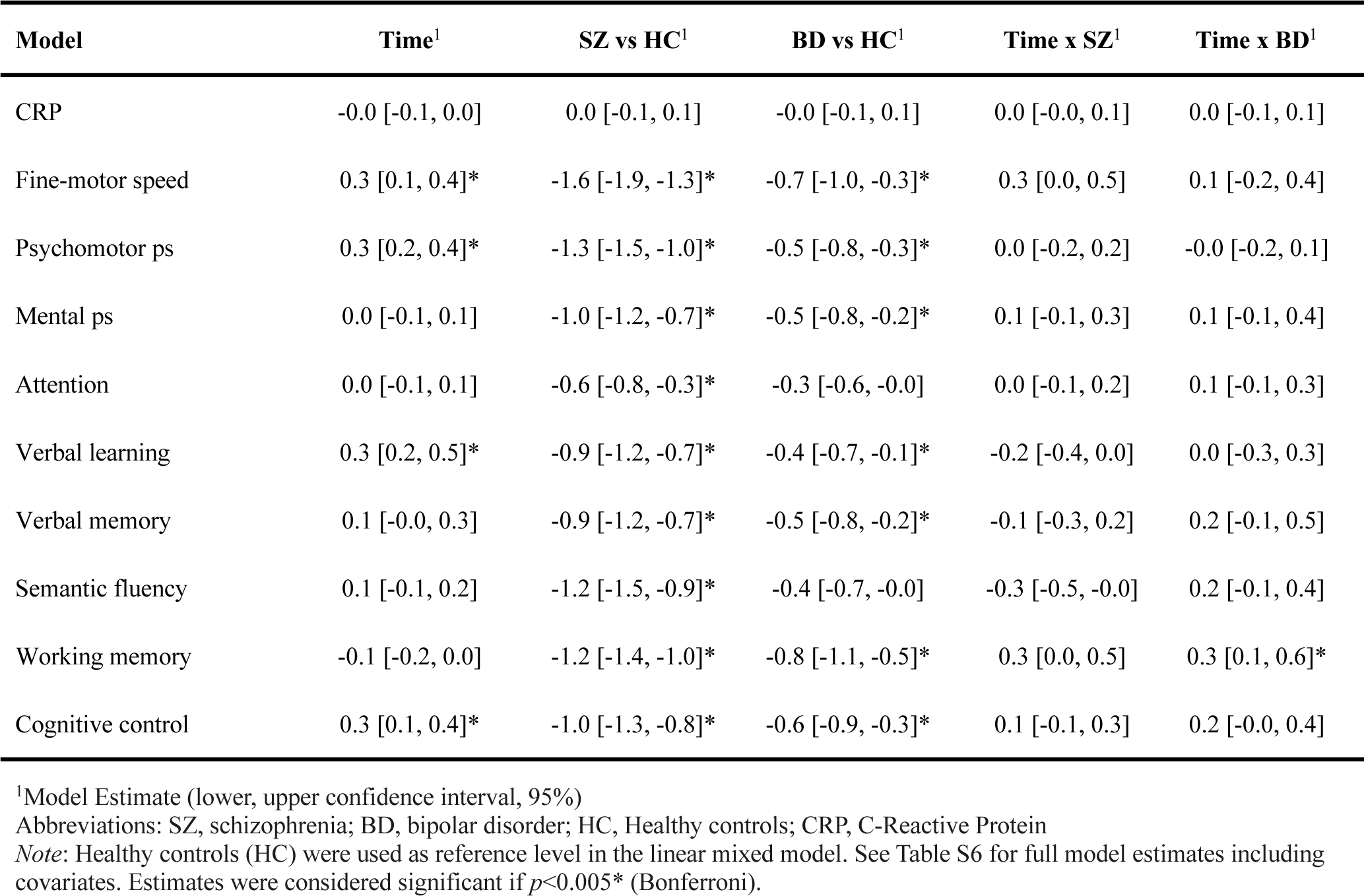
Results from mixed model analyses.

| Model | Time<sup>1</sup> | SZ vs HC<sup>1</sup> | BD vs HC<sup>1</sup> | Time x SZ<sup>1</sup> | Time x BD<sup>1</sup> |
| --- | --- | --- | --- | --- | --- |
| CRP | -0.0 [-0.1, 0.0] | 0.0 [-0.1, 0.1] | -0.0 [-0.1, 0.1] | 0.0 [-0.0, 0.1] | 0.0 [-0.1, 0.1] |
| Fine-motor speed | 0.3 [0.1, 0.4]* | -1.6 [-1.9, -1.3]* | -0.7 [-1.0, -0.3]* | 0.3 [0.0, 0.5] | 0.1 [-0.2, 0.4] |
| Psychomotor ps | 0.3 [0.2, 0.4]* | -1.3 [-1.5, -1.0]* | -0.5 [-0.8, -0.3]* | 0.0 [-0.2, 0.2] | -0.0 [-0.2, 0.1] |
| Mental ps | 0.0 [-0.1, 0.1] | -1.0 [-1.2, -0.7]* | -0.5 [-0.8, -0.2]* | 0.1 [-0.1, 0.3] | 0.1 [-0.1, 0.4] |
| Attention | 0.0 [-0.1, 0.1] | -0.6 [-0.8, -0.3]* | -0.3 [-0.6, -0.0] | 0.0 [-0.1, 0.2] | 0.1 [-0.1, 0.3] |
| Verbal learning | 0.3 [0.2, 0.5]* | -0.9 [-1.2, -0.7]* | -0.4 [-0.7, -0.1]* | -0.2 [-0.4, 0.0] | 0.0 [-0.3, 0.3] |
| Verbal memory | 0.1 [-0.0, 0.3] | -0.9 [-1.2, -0.7]* | -0.5 [-0.8, -0.2]* | -0.1 [-0.3, 0.2] | 0.2 [-0.1, 0.5] |
| Semantic fluency | 0.1 [-0.1, 0.2] | -1.2 [-1.5, -0.9]* | -0.4 [-0.7, -0.0] | -0.3 [-0.5, -0.0] | 0.2 [-0.1, 0.4] |
| Working memory | -0.1 [-0.2, 0.0] | -1.2 [-1.4, -1.0]* | -0.8 [-1.1, -0.5]* | 0.3 [0.0, 0.5] | 0.3 [0.1, 0.6]* |
| Cognitive control | 0.3 [0.1, 0.4]* | -1.0 [-1.3, -0.8]* | -0.6 [-0.9, -0.3]* | 0.1 [-0.1, 0.3] | 0.2 [-0.0, 0.4] |

<sup>1</sup>Model Estimate (lower, upper confidence interval, 95%)
Abbreviations: SZ, schizophrenia; BD, bipolar disorder; HC, Healthy controls; CRP, C-Reactive Protein
*Note:* Healthy controls (HC) were used as reference level in the linear mixed model. See Table S6 for full model estimates including covariates. Estimates were considered significant if $p < 0.005^*$ (Bonferroni).

### Subgroups based on inflammation and cognition

Evaluation of hierarchical clustering on CRP and the cognitive composite score revealed a 2-cluster solution to be optimal, with a favourable agglomerative coefficient (0.99) when using Ward’s linkage method (Fig. S3). The simulation procedure resulted in a significant silhouette index (*p*<0.001), rejecting the null hypothesis that the data comes from a single Gaussian distribution (Fig. S4). The cluster assignment was robust for both clusters following bootstrapping, with 81% (cluster 1) and 74% (cluster 2) overlap. As seen in Fig. 2A, the first cluster captured a subgroup (*n*=209, SZ=30 [25%], BD=45 [52%], HC=134 [62%]) characterized by a higher proportion of HC, lower inflammation and higher cognition (see Table 3), compared to the second subgroup (*n*=215, SZ=91 [75%], BD=42 [48%], HC=82 [38%]) which had a larger proportion of the SZ group, higher inflammation and lower cognition (chi square *p*<0.001, *d*=0.5-1.9). We additionally performed hierarchical clustering on the SMI group alone and found that the same inflammation-cognition pattern emerged, albeit characterized by even higher CRP levels and lower composite score in the higher inflammation – lower cognition subgroup, which also included predominantly SZ (Table S7).

**Figure 2.**
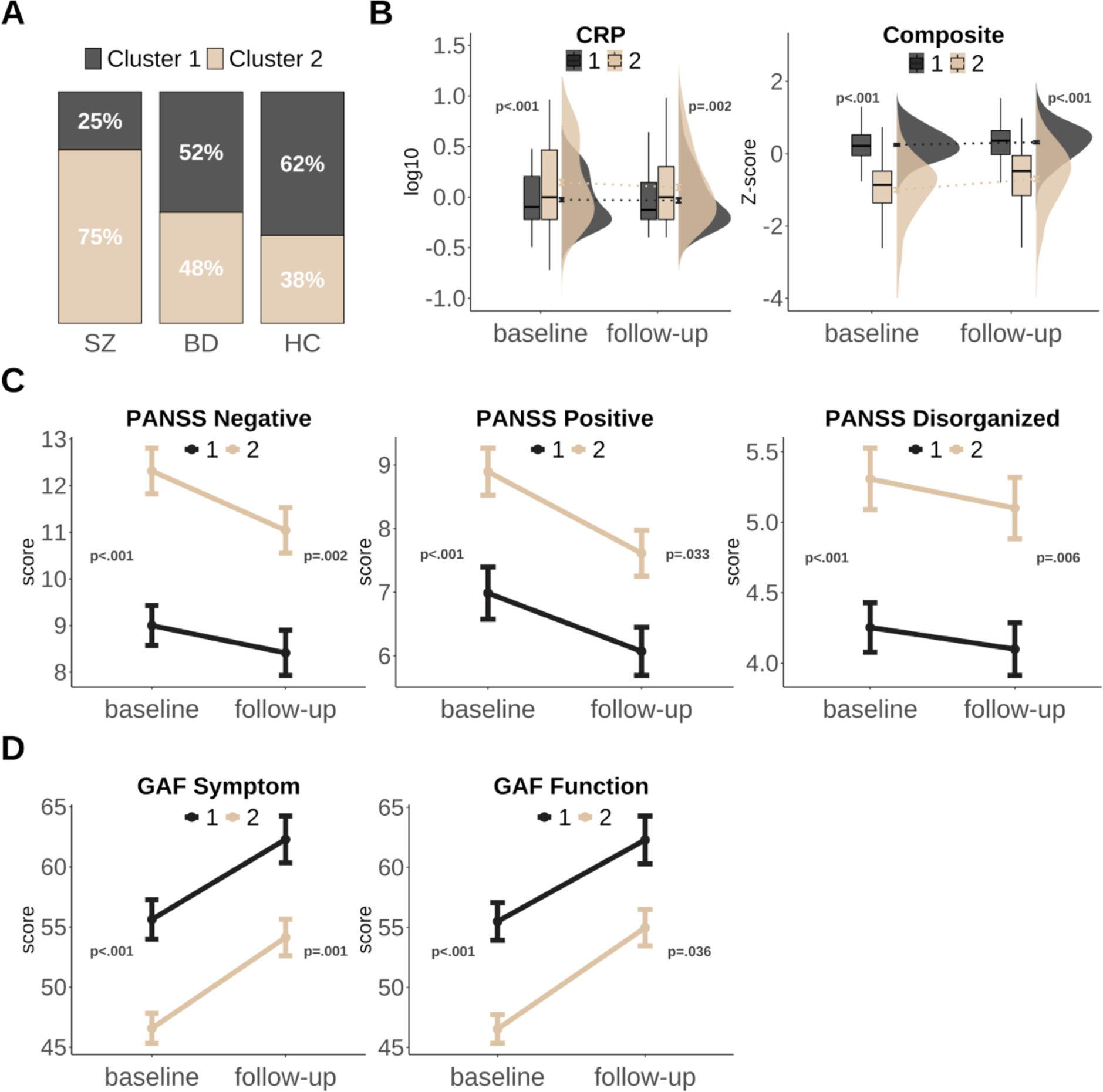
Inflammatory-cognitive subgroups at baseline and follow-up. Panel A shows the distribution of SZ, BD and HC in each cluster/subgroup. Panel B shows cluster differences in CRP (log10 transformed) and the cognitive composite score (Z-scores) at baseline and follow-up, with boxplots (interquartile range separated by median line), density plot (kernel density estimate) and line from mean scores (error bars: ±SEM). Panel C shows PANSS factors that were significantly different across clusters at both time points, and panel D shows differences across level of functioning (GAF symptom and GAF Function). Panel C and D is SMI only (line from mean, error bars: ±SEM).

**Table 3.** Subgroup comparisons on sample and clinical characteristics at baseline and follow-up.

| Characteristic | Subgroup 1<br>N = 209<sup>1</sup> | Subgroup 2<br>N = 215<sup>1</sup> | CI<sup>2</sup> | *p*<sup>3</sup> | Subgroup 1<br>N = 209<sup>1</sup> | Subgroup 2<br>N = 215<sup>1</sup> | CI<sup>2</sup> | *p*<sup>3</sup> |
| --- | --- | --- | --- | --- | --- | --- | --- | --- |
| *Baseline* |  |  |  |  | *Follow-up* |  |  |  |
| Age | 31.2 (8.8) | 29.7 (9.4) | [-0.3, 3.2] | ns | - | - | - | - |
| Sex (female %) | 114 (55) | 91 (42) | - | ns | - | - | - | - |
| Education (years) | 14.4 (2.2) | 13.2 (2.4) | [0.8, 1.6] | **<0.001** | - | - | - | - |
| WASI IQ (2-subtests) | 115.8 (9.0) | 104.8 (13.1) | [8.8, 13] | **<0.001** | - | - | - | - |
| BMI (kg/m<sup>2</sup>) | 24.2 (4.0) | 24.9 (4.0) | [-1.5, 0.0] | ns | 24.1 (4.3) | 25.3 (3.9) | [-2.5, 0.0] | ns |
| Composite, cognition | 0.3 (0.4) | -1.0 (0.8) | [1.1, 1.4] | **<0.001** | 0.3 (0.5) | -0.7 (1.0) | [0.9, 1.2] | **<0.001** |
| CRP | 1.1 (0.7) | 2.1 (2.0) | [-1.3, -0.7] | **<0.001** | 1.2 (1.1) | 1.8 (1.8) | [-0.9, -0.2] | **0.002** |
| **SMI only** |  |  |  |  |  |  |  |  |
| PANSS Negative | 9.0 (3.7) | 12.3 (5.6) | [-4.6, -2.0] | **<0.001** | 8.4 (4.1) | 11.0 (5.3) | [-4.0, -1.3] | **0.002** |
| PANSS Positive | 7.0 (3.6) | 8.9 (4.3) | [-3.0, -0.8] | **<0.001** | 6.1 (3.2) | 7.6 (3.9) | [-2.6, -0.5] | **0.033** |
| PANSS Disorganized | 4.3 (1.5) | 5.3 (2.5) | [-1.6, -0.5] | **<0.001** | 4.1 (1.6) | 5.1 (2.4) | [-1.6, -0.4] | **0.006** |
| PANSS Excited | 5.2 (1.6) | 5.1 (1.5) | [-0.3, 0.5] | ns | 5.0 (1.8) | 4.8 (1.3) | [-0.3, 0.6] | ns |
| PANSS Depressed | 8.6 (3.0) | 8.3 (3.0) | [-0.5, 1.2] | ns | 6.7 (2.9) | 7.1 (3.0) | [-1.3, 0.4] | ns |
| YMRS | 3.9 (4.1) | 4.9 (5.5) | [-2.3, 0.4] | ns | 3.1 (4.5) | 4.1 (4.5) | [-2.4, 0.3] | ns |
| GAF Symptom | 55.6 (14.2) | 46.6 (14.4) | [5.0, 13] | **<0.001** | 62.3 (15.9) | 54.1 (16.6) | [3.3, 13] | **0.011** |
| GAF Function | 55.5 (13.6) | 46.5 (13.7) | [5.1, 13] | **<0.001** | 62.3 (16.2) | 55.0 (16.5) | [2.4, 12] | **0.036** |
| DUI | 105.6 (193.3) | 102.0 (199.2) | [-64, 71] | ns | - | - | - | - |
| Total, DDD | 1.5 (1.0) | 1.9 (1.1) | [-0.7, 0.0] | ns | 2.8 (1.1) | 1.7 (0.9) | [-0.3, 2.3] | ns |

<sup>1</sup>Mean (SD); n (%)<sup>2</sup>CI = Confidence Interval, 95%<sup>3</sup>Welch Two Sample t-test; Pearson's Chi-squared test (Bonferroni corrected *p*-values)
Abbreviations: WASI, Wechsler Abbreviated Scale of Intelligence; CRP, C-reactive Protein; BMI, body mass index; PANSS, Positive and Negative Syndrome Scale; YMRS, Young Mania Rating Scale; GAF, Global Assessment of Functioning scale; DUI, Days of untreated illness; DDD, defined daily dosage.
Note: Subgroup 1 = lower inflammation – higher cognition; Subgroup 2 = higher inflammation – lower cognition

### Characteristics of inflammatory-cognitive subgroups at baseline and follow-up

As seen in Fig. 2B, the subgroup pattern was consistent over time, with higher inflammation and lower cognition in the second subgroup relative to the first also at follow-up (*d*=0.4-1.3, Table 3). Relative to the first subgroup, the higher inflammation – lower cognition subgroup had shorter education and lower IQ (all *p*<0.001, *d*=0.5-0.9), but they did not differ in age, sex, or BMI. The higher inflammation – lower cognition subgroup had lower scores on all cognitive domains both at baseline (*d*=0.8-1.4) and follow-up (*d*=0.5-1.1) compared to the lower inflammation – higher cognition subgroup (all *p*<0.001, Fig. S5). Compared to the other subgroup, participants with SMI in the higher inflammation – lower cognition subgroup had more positive, negative, and disorganised symptoms (Fig. 2C), as well as lower functioning (GAF_S_ and GAF_F_; Fig. 2D), at both time points (*d*_baseline_=0.5-0.7, *d*_follow-up_=0.4-0.5). Regardless of group, there was a significant improvement in the cognitive composite score (*p*<0.001), and all symptoms and functioning scores (all *p*<0.001), except for disorganized symptoms which remained stable. The level of CRP however, remained stable (*p*=0.623). Analysis of change scores revealed a slightly higher gain in cognitive performance from baseline to follow-up in the second subgroup compared to the first (*p*<0.001, Wilcoxon effect size, *r*=0.2(small)). There was no difference in change scores between the subgroups on any symptoms or functioning measures.

## Discussion

This study evaluated the longitudinal course of inflammation and cognition in a large sample of first treatment SZ and BD, and a HC cohort. While there were case-control differences in CRP at baseline or follow-up, we identified two transdiagnostic inflammatory-cognitive subgroups with differing levels of clinical and functional characteristics. The higher inflammation – lower cognition subgroup (predominantly SZ) had more symptoms and lower functioning at both time-points, compared to the lower inflammation – higher cognition subgroup. While inflammation, cognition, symptoms, and functioning remained stable or improved over time for both subgroups, the higher inflammation – lower cognition group still scored well below the other subgroup at follow-up. The fact that SZ, BD, and HC were represented in both subgroups shows that heterogeneity is characteristic for both inflammation and cognition. Our findings suggest transdiagnostic inflammatory-cognitive subgroups that are stable across time. This indicates that the inflammatory-cognitive association may be more trait-than state-related.

The main finding is that inflammatory-cognitive subgroups based on CRP as a measure of inflammation and a cognitive composite score, is stable over one year in first treatment SMI and HC. These findings also confirm the inflammatory-cognitive subgroup pattern that we previously identified using broad panels of inflammatory and immune-related markers and cognitive domains (Sæther et al., 2023, 2024). Importantly, while cognition, symptoms, and level of functioning generally improved over the first year of treatment for SMI participants, we observed stable differences between the subgroups at both time-points, with the higher inflammation – lower cognition subgroup having worse cognition, higher inflammation, more symptoms, and lower functioning. Results from clinical trials suggest that add-on anti-inflammatory treatments are more effective in SMI patients exhibiting higher inflammation (Jeppesen et al., 2020; Nettis et al., 2021). Similarly, cognitive remediation may be particularly efficacious for patients with significant cognitive impairments, although those with milder impairments also benefit (Vita et al., 2021; Wykes et al., 2011). Given the between-subgroup stability in characteristics (inflammatory, cognitive, clinical) over time, these subgroups could be ideal candidates for personalized interventions. For the more impaired subgroup this could include cognitive remediation combined with anti-inflammatory add-on treatments, as the latter may also have beneficial effects on cognition (Jeppesen et al., 2020). One could speculate that HC in the impaired subgroup constitute a vulnerable group, particularly since low-grade inflammation is also a risk factor in the general population for developing autoimmune-, cardiovascular- and neurodegenerative disease (Furman et al., 2019). It is worth noting that 36% of the SMI group showed a similar pattern to the HC group (i.e. those in the lower inflammation – higher cognition group) with a positive clinical trajectory. This group may benefit from other interventions that should also focus on cognitive strengths (Allott et al., 2020).

As shown in previous studies with overlapping samples (Demmo et al., 2017; Engen et al., 2019; Flaaten et al., 2023b, 2023a, 2022; Haatveit et al., 2015), our analyses comparing diagnostic status show domain-specific stability or improvement in cognitive functioning from baseline to follow-up. This is in line with longitudinal findings in SMI from other groups (Bora and Özerdem, 2017; Catalan et al., 2024; Torgalsbøen et al., 2023). A similar course of improvement in both SMI and HC may indicate practice effects, which is known for some of the cognitive tests used in this study (Beglinger et al., 2005). In terms of subgroups, we observed that while the higher inflammation – lower cognition subgroup had a slight improvement in cognition, they still performed significantly lower than the lower inflammation – higher cognition subgroup at follow-up. Sustained cognitive impairment is strongly associated with poor functional outcomes (Cowman et al., 2021), underscoring the need to develop and implement effective treatments for cognitive impairment in SMI.

Our data did not suggest case-control differences in CRP levels at baseline or follow-up. While meta-analyses have reported consistent evidence of elevated CRP in SMI compared to HC, it may be higher during acute manic or psychotic episodes (Fernandes et al., 2016; Fond et al., 2018; Halstead et al., 2023; Lestra et al., 2022). However, participants in the TOP-study have been evaluated in euthymic/milder symptom states. We accounted for age, sex and BMI which has been shown to attenuate CRP findings on psychiatric symptoms (Figueroa-Hall et al., 2022). These covariates are not always included in studies reported by meta-analyses (Fernandes et al., 2016; Fond et al., 2018; Halstead et al., 2023; Lestra et al., 2022). Further, inflammatory markers in SMI are typically in the smaller effect size range (Carvalho et al., 2020; Miller and Goldsmith, 2020). This may pose a challenge for detecting case-control differences, as only a subset of individuals with SMI show elevated levels of inflammation (Bishop et al., 2022; Miller and Goldsmith, 2019). Nonetheless, the higher inflammation – lower cognition subgroup suggests some interaction with CRP and cognition, particularly in SZ participants that were overrepresented in this subgroup. This also aligns with previous findings that individuals with SMI in high-inflammatory subgroups have lower cognitive performance (Fillman et al., 2016; Lizano et al., 2023, 2020). Our findings suggest that there are trait-related cognitive-immune subgroups in SMI, which seems independent of state dependent fluctuations of immune markers.

There are some limitations to consider. We cannot exclude the possibility that more complex subgroup patterns would emerge with larger samples and/or more inflammatory markers, as suggested by recent machine learning approaches (Lalousis et al., 2023). While CRP is an inexpensive and accessible marker of systemic inflammation, it cannot provide further insight about specific inflammatory pathways or mechanisms that might be related to cognitive impairment. Future studies should therefore include a broader spectrum of inflammatory and immune-related markers in longitudinal designs. Although we found stability in inflammatory-cognitive subgroups over time, the study was unable to establish whether inflammation and lower cognition simply co-occurs or has a causal relationship. There are also potentially other factors that might influence both cognition and inflammation that were not accounted for in this study. Strengths of this study lie in the longitudinal design, the large sample of first treatment SMI and the inclusion of HC, as well as the robust evaluation of the clustering solution with stability analyses. However, our findings should be replicated using independent samples.

## Conclusion

Results from our study suggest that transdiagnostic inflammatory-cognitive subgroups defined at baseline are stable over time. Individuals with SMI in the higher inflammation – lower cognition subgroup had sustained symptoms and lower functioning, suggesting a specific phenotype that may benefit from personalized treatments targeting both inflammation and cognition.

## Data Availability

Some data in the present study may be made available upon reasonable request.

## Acknowledgements

We wish to thank all participants for their valuable contribution to the TOP-study and staff for their contribution to data collection and curation. Funding for this project was provided by the South-Eastern Norway Regional Health Authority (grant #2020089, #2023031) and Research Council of Norway (#223273, #326813). We wish to acknowledge Sigma2 (the National Infrastructure for High Performance Computing and Data Storage in Norway), Services for Sensitive Data (TSD) at the University of Oslo, and the Department of Medical Biochemistry at Oslo University Hospital.

