## Supplementary material for "Longitudinal course of inflammatory-cognitive subgroups across first treatment severe mental illness and healthy controls": Supp_Figs

**Supplementary Figures**

**1. Correlation between CRP and cognitive domains at baseline and follow-up**

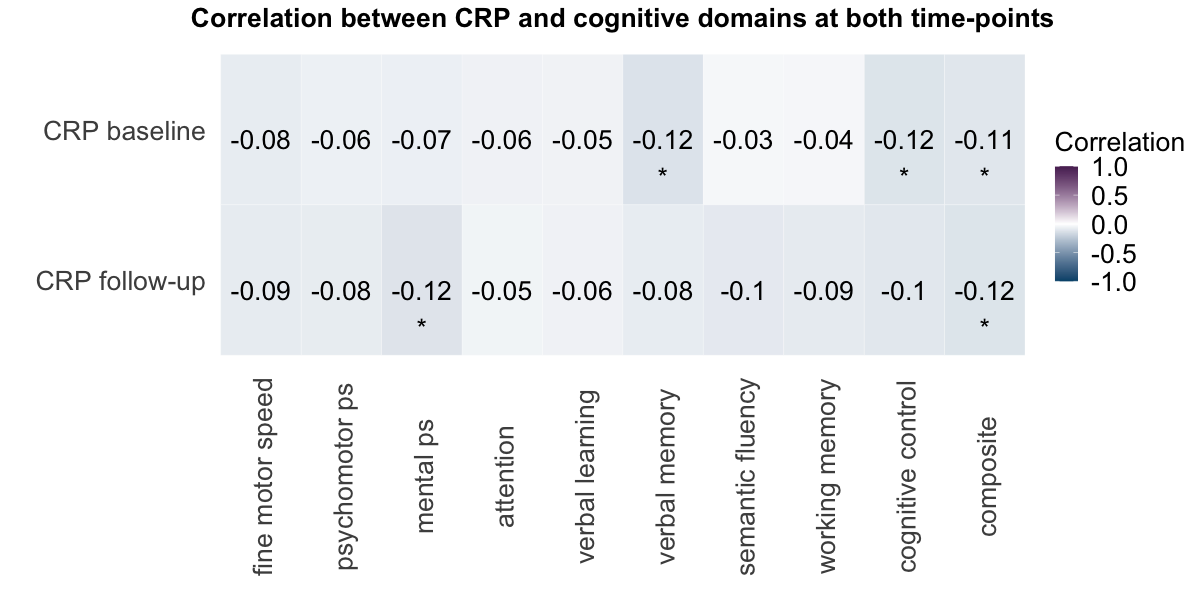

**Supplementary Fig. 1** Correlation (Pearson) of CRP (log10 transformed) with all cognitive domains and a composite score in A) at baseline, and B) at follow-up.

**2. Cognition at baseline and follow-up comparing diagnostic status**

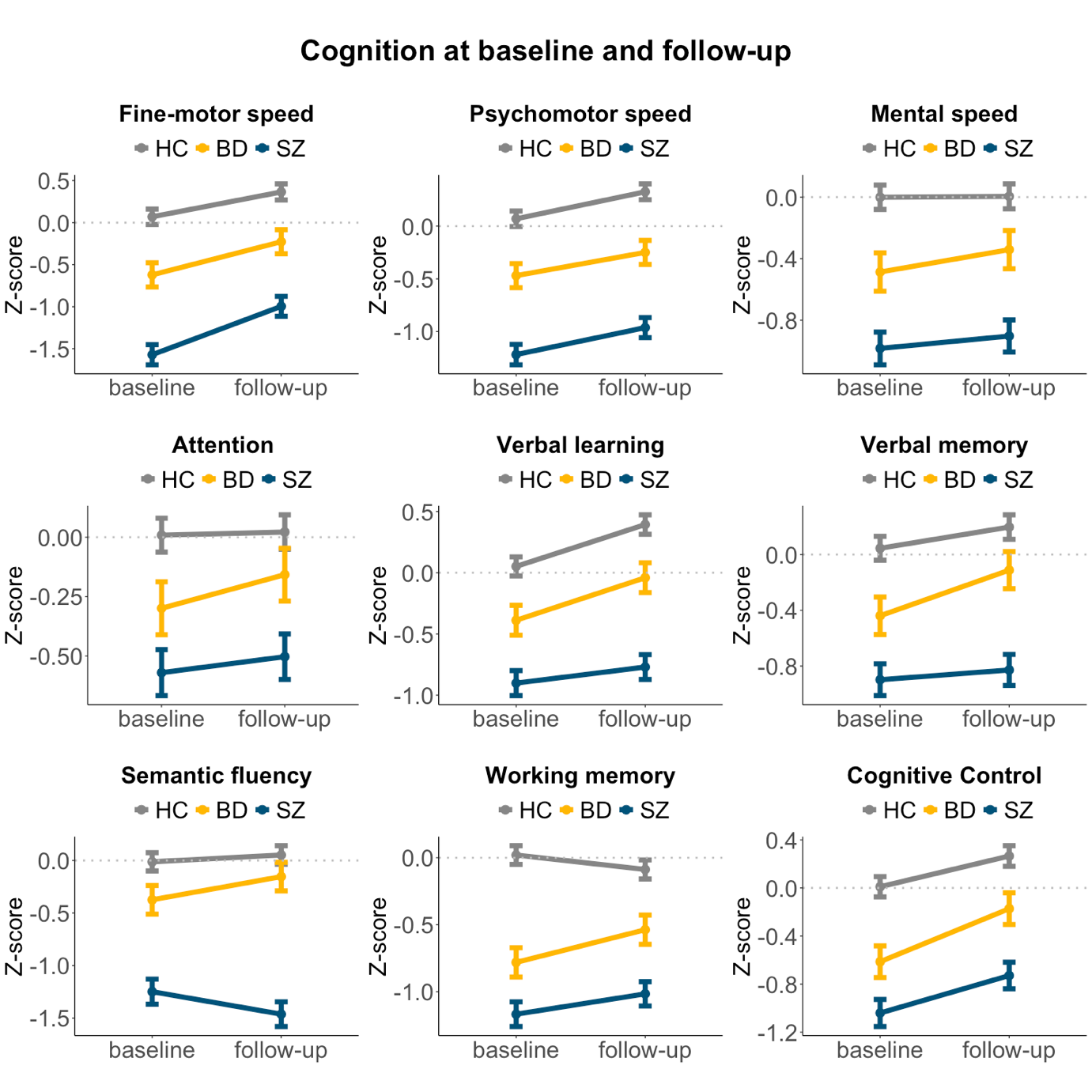

**Supplementary Fig. 2** Estimated marginal means (including error bars: ±SEM) for cognitive domains at baseline and follow-up between SZ, BD, and HC.

**3. Hierarchical clustering: Evaluation of optimal number of clusters**

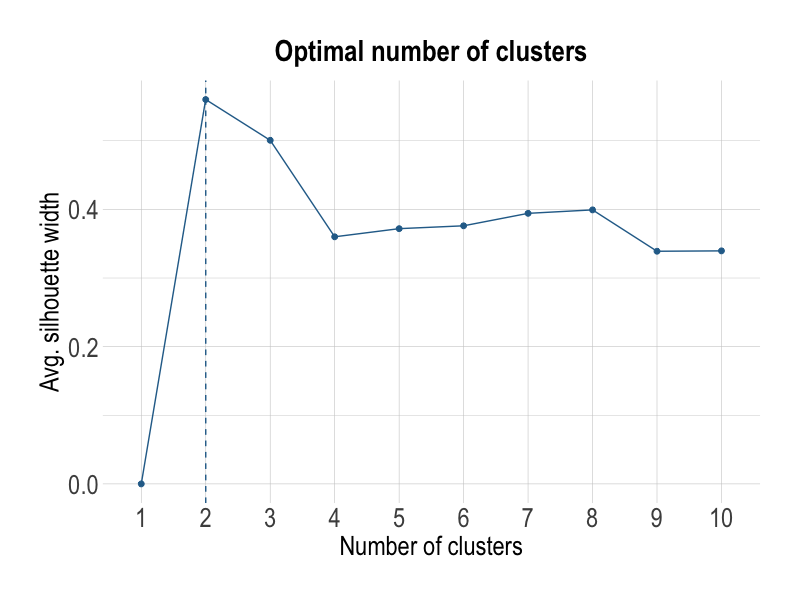
**Supplementary Fig. 3** Evaluation of n clusters for hierarchical clustering of baseline global cognitive score and CRP using the average silhouette index. The average silhouette index was maximized for a 2-cluster solution.

**4. Hierarchical clustering: Significance test**

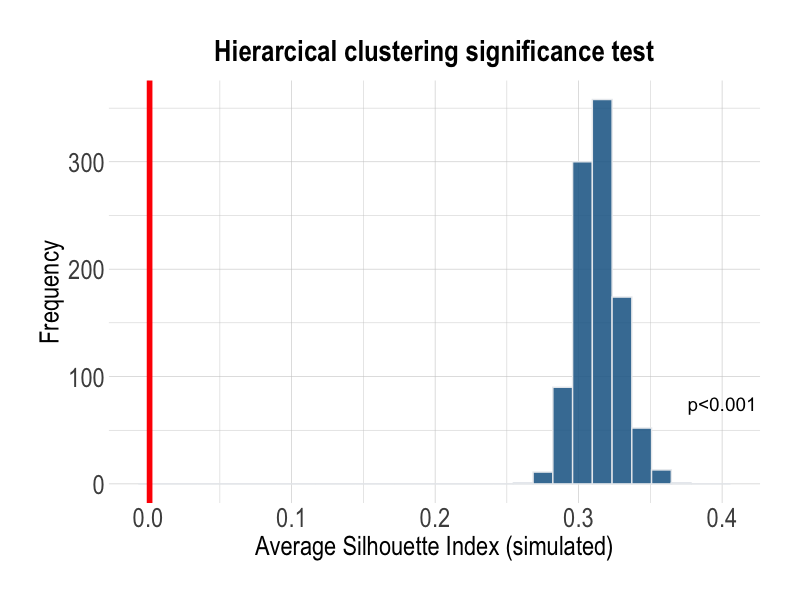

**Supplementary Fig. 4** Shows null distribution and results from significance test of the average silhouette index. A significant result (*p*<0.001) indicates we can reject the null hypothesis (i.e. data comes from a single normal Gaussian distribution).

**5. Hierarchical clustering: Comparison of clusters on cognitive performance at baseline and follow-up**

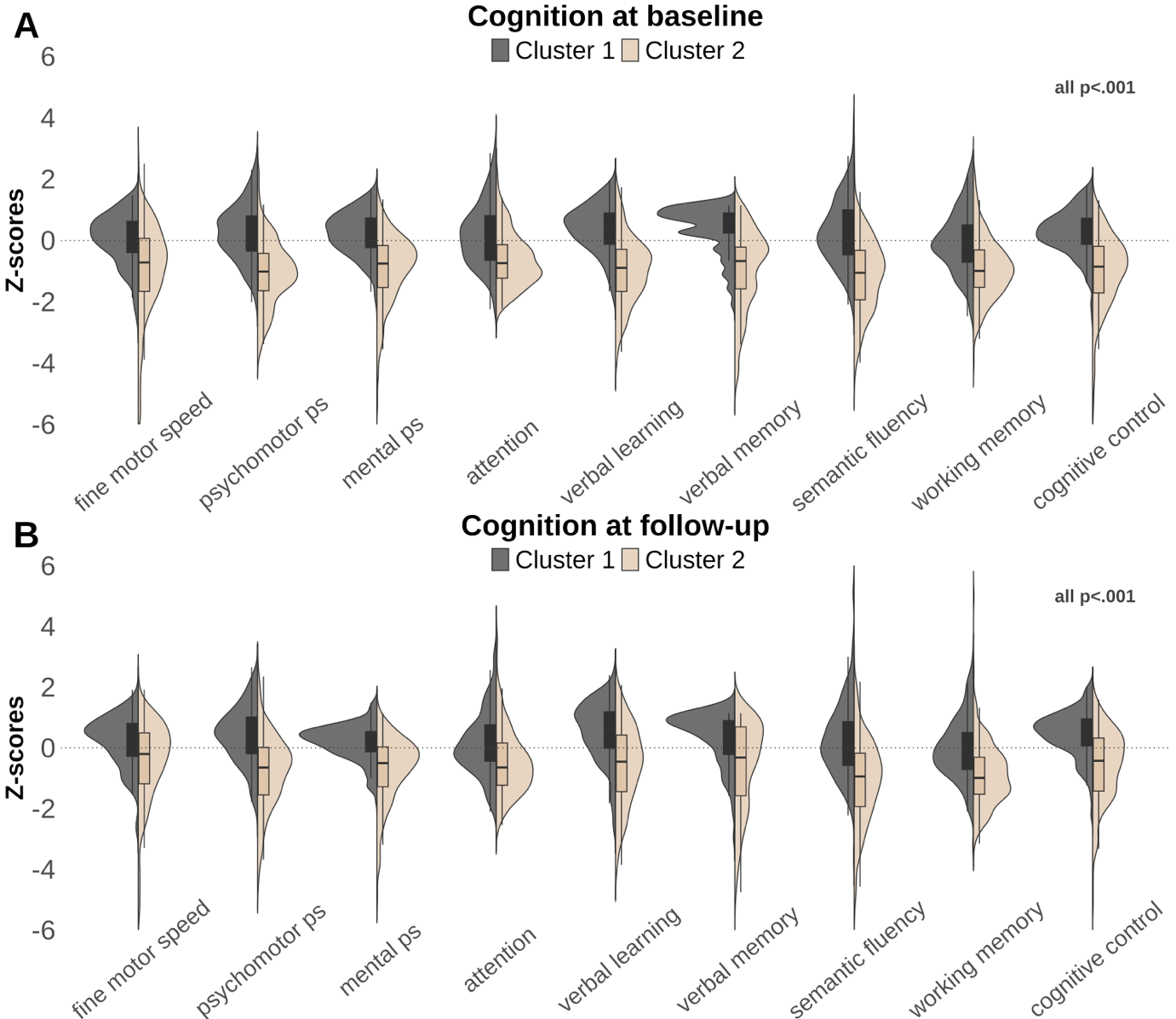

**Supplementary Fig. 5** Shows comparisons between clusters (based on CRP and a composite score at baseline) across cognitive domains at baseline and follow-up.
