## Supplementary material for "Longitudinal course of inflammatory-cognitive subgroups across first treatment severe mental illness and healthy controls": Supp_Methods

**Supplementary Methods**

1. **R-packages used for visualization and main statistical analyses**

- For visualization the R-package “ggplot2” was applied (Wickham, 2016).
- For permutation-based t-tests (sample & clinical characteristics) the R-package “rcompanion” (Mangiafico, 2022) was used.
- Clustering was performed using R-packages “cluster” (Maechler et al., 2022), “dendextend” (Galili, 2015) and “factoextra” (Kassambara and Mundt, 2020).
- Clustering stability was assessed using the R-package “fpc” (flexible procedures for clustering) (Hennig, 2020).
- Linear mixed models were run using “lme4" (Bates et al., 2015), “lmerTest” (Kuznetsova et al., 2017) and model comparisons with “emmeans” (Lenth et al., 2018).
- Tables were automatically generated using “gtsummary” (Sjoberg et al., 2021).
