## Supplementary material for "Longitudinal course of inflammatory-cognitive subgroups across first treatment severe mental illness and healthy controls": Supp_Tables

**Supplementary Tables**

| **Supplementary Table 1.** Somatic medication use by SMI group | | |
| --- | --- | --- |
| **Somatic medications** | **SZ**  **(n=133)** | **BD (n=88)** |
| Anti-inflammatory/immunomodulatory, N (%) | 4 (3.0) | 0 (0.0) |
| Antidiabetics, N (%) | 2 (1.5) | 1 (1.1) |
| Cardiovascular/lipid modifying, N (%) | 3 (2.2) | 0 (0.0) |
| Antihistamines, N (%) | 5 (3.7) | 4 (4.5) |
| Gastrointestinal agents, N (%) | 2 (1.5) | 1 (1.1) |
| *Other, N (%) | 4 (3.0) | 8 (9.1) |
| *** Other includes vitamins, minerals, analgetics, thyroid agents, pulmonary agents, urological agents, musculoskeletal agents, contraceptives, sex hormones, anxiolytics, anti-inflammatory agents (local administrative agents, i.e. ointments/inhalators), hematological agents, parenteral nutrition agents, substance dependency agents and mucolytic agents.  Abbreviations: schizophrenia (SZ), bipolar disorder (BD), healthy controls (HC) | | |

| **Supplementary Table 2.** Overview of cognitive domains and corresponding tests from test battery I and II | |
| --- | --- |
| ***Domain*/Test** | **Test battery** |
| ***Psychomotor processing speed*** |  |
| Symbol Coding (WAIS-III) | I |
| BACS Symbol Coding (MCCB) | II |
| ***Verbal learning*** |  |
| Total recall (CVLT-II) | I |
| Total recall (HVLT-R, MCCB) | II |
| ***Verbal memory*** |  |
| Long-delay free recall (CVLT-II) | I |
| Delayed recall (HVLT-R, MCCB) | II |
| ***Semantic fluency*** |  |
| Category fluency (D-KEFS) | I |
| Category fluency (MCCB) | II |
| ***Working memory*** |  |
| Letter Number Sequencing (WAIS-III) | I |
| Letter Number Sequencing (MCCB) | II |
| ***Fine motor speed*** |  |
| Grooved Pegboard test (Halstead-Reitan) | I & II |
| ***Attention*** |  |
| Digit Span Forward (WAIS-III) | I & II |
| ***Mental processing speed*** |  |
| Color naming+Color reading (D-KEFS) | I & II |
| ***Cognitive control*** |  |
| Inhibition+Inhibition switching (D-KEFS) | I & II |
| Abbreviations: WAIS, Wechsler Abbreviated Scale of Intelligence; MCCB, the MATRIX Consensus Cognitive Battery; CVLT, California Verbal Learning Test; HVLT-R, Hopkins Verbal Learning Test Revised; D-KEFS, Delis-Kaplan Executive Function System. | |

| **Supplementary Table 3.** Overview of number of observations in each group for inflammatory marker and cognitive domains at baseline and follow-up | | | | | | |
| --- | --- | --- | --- | --- | --- | --- |
|  | **Baseline** | | | **Follow-up** | | |
| **Characteristic** | **HC** | **BD** | **SZ** | **HC** | **BD** | **SZ** |
| CRP | 216 | 87 | 127 | 174 | 83 | 99 |
| Fine-motor speed | 211 | 81 | 126 | 190 | 83 | 126 |
| Psychomotor speed | 219 | 88 | 132 | 219 | 87 | 132 |
| Mental speed | 219 | 87 | 125 | 219 | 86 | 128 |
| Attention | 220 | 88 | 124 | 220 | 87 | 123 |
| Verbal learning | 220 | 87 | 133 | 219 | 88 | 132 |
| Verbal memory | 220 | 86 | 126 | 212 | 88 | 131 |
| Semantic fluency | 219 | 88 | 120 | 219 | 88 | 121 |
| Working memory | 219 | 88 | 131 | 218 | 87 | 132 |
| Cognitive control | 217 | 87 | 125 | 219 | 86 | 128 |
| Abbreviations: SZ, schizophrenia; BD, bipolar disorder; HC, Healthy controls; CRP, C-Reactive Protein. | | | | | | |

| **Supplementary Table 4.** Descriptive statistics for inflammatory marker and cognitive domains from baseline to follow-up | | | | | | |
| --- | --- | --- | --- | --- | --- | --- |
|  | **Baseline**^1^ | | | **Follow-up**^1^ | | |
| **Characteristic** | **HC**  (N = 220)^2^ | **BD**  (N = 88)^2^ | **SZ**  (N = 133)^2^ | **HC**  (N = 220)^2^ | **BD**  (N = 88)^2^ | **SZ**  (N = 133)^2^ |
| CRP | 1.63 (1.63) | 1.59 (1.58) | 1.56 (1.43) | 1.47 (1.45) | 1.41 (1.43) | 1.67 (1.89) |
| Fine-motor speed | 0.00 (0.90) | -0.53 (1.59) | -1.53 (1.95) | 0.28 (0.76) | -0.16 (1.22) | -0.97 (1.81) |
| Psychomotor speed | 0.00 (1.00) | -0.37 (1.15) | -1.18 (1.17) | 0.23 (0.98) | -0.18 (1.18) | -0.95 (1.33) |
| Mental speed | 0.00 (0.91) | -0.46 (0.98) | -1.06 (1.53) | 0.01 (0.75) | -0.32 (0.92) | -0.96 (1.79) |
| Attention | 0.00 (1.00) | -0.30 (1.05) | -0.54 (0.99) | 0.01 (1.03) | -0.17 (1.08) | -0.48 (1.12) |
| Verbal learning | 0.00 (1.00) | -0.31 (1.22) | -0.86 (1.20) | 0.33 (1.03) | 0.02 (1.27) | -0.75 (1.42) |
| Verbal memory | 0.00 (1.00) | -0.36 (1.35) | -0.83 (1.47) | 0.13 (1.10) | -0.06 (1.16) | -0.82 (1.63) |
| Semantic fluency | 0.00 (1.00) | -0.38 (1.27) | -1.28 (1.50) | 0.07 (1.15) | -0.15 (1.42) | -1.52 (1.51) |
| Working memory | 0.00 (1.00) | -0.78 (0.95) | -1.11 (1.01) | -0.12 (1.04) | -0.54 (1.10) | -0.97 (1.03) |
| Cognitive control | 0.00 (0.88) | -0.58 (1.21) | -1.09 (1.79) | 0.25 (0.86) | -0.11 (1.00) | -0.78 (1.68) |
| ^1^Mean (SD)  ^2^The number of observations for the inflammatory marker and cognitive domain varied, see Table X for overview  Abbreviations: SZ, schizophrenia; BD, bipolar disorder; HC, Health control; CRP, C-Reactive Protein. | | | | | | |

| **Supplementary Table 5.** Sample/clinical characteristics at follow-up | | | |
| --- | --- | --- | --- |
| **Characteristics** | **SZ** N = 133^1^ | **BD** N = 88^1^ | ***p*-value**^2^ |
| BMI (kg/m²) | 25.40 (4.43) | 24.44 (3.54) | ns |
| PANSS Negative | 11.95 (5.61) | 7.59 (2.40) | <0.001 |
| PANSS Positive | 8.56 (4.09) | 4.93 (1.39) | <0.001 |
| PANSS Disorganized | 5.38 (2.41) | 3.90 (1.38) | <0.001 |
| PANSS Excited | 4.99 (1.49) | 4.57 (1.40) | 0.013 |
| PANSS Depressed | 7.25 (2.94) | 6.61 (2.95) | ns |
| YMRS | 4.37 (4.70) | 2.48 (3.86) | 0.002 |
| GAF Symptom | 49.75 (15.30) | 67.18 (12.45) | <0.001 |
| GAF Function | 52.27 (15.30) | 65.36 (15.49) | <0.001 |
| Antipsychotics, DDD | 1.26 (0.98) | 2.33 (10.05) | <0.001 |
| Antidepressants, DDD | 1.29 (0.67) | 1.04 (0.76) | ns |
| Antiepileptics, DDD | 0.40 (0.21) | 0.91 (0.37) | <0.001 |
| Lithium, DDD | 0.88 (0.17) | 1.16 (0.52) | ns |
| Total, DDD | 3.05 (0.77) | 1.94 (1.02) | ns |
| ^1^Mean (SD)  ^2^Wilcoxon rank sum test Abbreviations: SZ, schizophrenia; BD, bipolar disorder; BMI, body mass index; PANSS, Positive and Negative Syndrome Scale; YMRS, Young Mania Rating Scale; GAF, Global Assessment of Functioning scale; DDD, defined daily dosage; ns, non-significant | | | |

| **Supplementary Table 6.** Estimates from mixed model analyses | | | | | | | |
| --- | --- | --- | --- | --- | --- | --- | --- |
| **Model** | **Parameter** | **Estimate** | **SE** | ***t*** | ***p*** | **CI lower** | **CI upper** |
| CRP | Intercept | -0.46 | 0.09 | -4.89 | **<0.001** | -0.65 | -0.28 |
|  | Time | -0.03 | 0.03 | -1.25 | 0.213 | -0.08 | 0.02 |
|  | Group: SZ | 0.01 | 0.04 | 0.18 | 0.858 | -0.07 | 0.08 |
|  | Group: BD | -0.02 | 0.04 | -0.57 | 0.567 | -0.11 | 0.06 |
|  | Sex: Male | -0.06 | 0.03 | -2.20 | 0.028 | -0.12 | -0.01 |
|  | Age | 0.00 | 0.00 | -0.98 | 0.328 | 0.00 | 0.00 |
|  | BMI | 0.02 | 0.00 | 7.02 | **<0.001** | 0.02 | 0.03 |
|  | Time x Group: SZ | 0.04 | 0.04 | 0.90 | 0.369 | -0.04 | 0.12 |
|  | Time x Group: BD | 0.02 | 0.05 | 0.47 | 0.642 | -0.07 | 0.11 |
| Fine-motor speed | Intercept | 1.08 | 0.26 | 4.20 | **<0.001** | 0.58 | 1.59 |
|  | Time | 0.30 | 0.07 | 4.05 | **<0.001** | 0.15 | 0.44 |
|  | Group: SZ | -1.64 | 0.15 | -10.61 | **<0.001** | -1.94 | -1.34 |
|  | Group: BD | -0.69 | 0.17 | -4.01 | **<0.001** | -1.03 | -0.35 |
|  | Sex: Male | -0.60 | 0.12 | -5.06 | **<0.001** | -0.83 | -0.37 |
|  | Age | -0.02 | 0.01 | -3.36 | **<0.001** | -0.04 | -0.01 |
|  | Time x Group: SZ | 0.28 | 0.12 | 2.41 | 0.016 | 0.05 | 0.51 |
|  | Time x Group: BD | 0.10 | 0.13 | 0.73 | 0.464 | -0.16 | 0.36 |
| Psychomotor speed | Intercept | 1.03 | 0.21 | 4.93 | **<0.001** | 0.62 | 1.43 |
|  | Time | 0.26 | 0.05 | 4.98 | **<0.001** | 0.16 | 0.36 |
|  | Group: SZ | -1.29 | 0.12 | -10.35 | **<0.001** | -1.53 | -1.05 |
|  | Group: BD | -0.54 | 0.14 | -3.92 | **<0.001** | -0.81 | -0.27 |
|  | Sex: Male | -0.53 | 0.10 | -5.53 | **<0.001** | -0.72 | -0.34 |
|  | Age | -0.02 | 0.01 | -4.00 | **<0.001** | -0.03 | -0.01 |
|  | Time x Group: SZ | 0.00 | 0.08 | 0.00 | 0.999 | -0.16 | 0.16 |
|  | Time x Group: BD | -0.04 | 0.10 | -0.37 | 0.710 | -0.22 | 0.15 |
| Mental speed | Intercept | 0.09 | 0.22 | 0.42 | 0.675 | -0.34 | 0.53 |
|  | Time | 0.01 | 0.06 | 0.10 | 0.924 | -0.11 | 0.12 |
|  | Group: SZ | -0.98 | 0.14 | -7.22 | **<0.001** | -1.25 | -0.72 |
|  | Group: BD | -0.49 | 0.15 | -3.26 | **0.001** | -0.78 | -0.19 |
|  | Sex: Male | -0.31 | 0.10 | -3.01 | **0.003** | -0.51 | -0.11 |
|  | Age | 0.00 | 0.01 | 0.32 | 0.746 | -0.01 | 0.01 |
|  | Time x Group: SZ | 0.07 | 0.10 | 0.75 | 0.455 | -0.12 | 0.27 |
|  | Time x Group: BD | 0.14 | 0.11 | 1.23 | 0.219 | -0.08 | 0.36 |
| Attention | Intercept | 0.06 | 0.20 | 0.32 | 0.752 | -0.32 | 0.45 |
|  | Time | 0.01 | 0.06 | 0.22 | 0.826 | -0.10 | 0.13 |
|  | Group: SZ | -0.58 | 0.12 | -4.70 | **<0.001** | -0.82 | -0.34 |
|  | Group: BD | -0.31 | 0.13 | -2.3 | 0.022 | -0.57 | -0.05 |
|  | Sex: Male | 0.14 | 0.09 | 1.56 | 0.119 | -0.04 | 0.32 |
|  | Age | 0 | 0.01 | -0.76 | 0.447 | -0.01 | 0.01 |
|  | Time x Group: SZ | 0.05 | 0.1 | 0.54 | 0.587 | -0.14 | 0.25 |
|  | Time x Group: BD | 0.13 | 0.11 | 1.15 | 0.250 | -0.09 | 0.35 |
| Verbal learning | Intercept | 0.8 | 0.21 | 3.79 | **<0.001** | 0.39 | 1.21 |
|  | Time | 0.34 | 0.07 | 4.69 | **<0.001** | 0.2 | 0.48 |
|  | Group: SZ | -0.95 | 0.13 | -7.24 | **<0.001** | -1.21 | -0.70 |
|  | Group: BD | -0.44 | 0.15 | -3 | **0.003** | -0.73 | -0.15 |
|  | Sex: Male | -0.4 | 0.1 | -4.19 | **<0.001** | -0.59 | -0.22 |
|  | Age | -0.02 | 0.01 | -3.14 | **0.002** | -0.03 | -0.01 |
|  | Time x Group: SZ | -0.21 | 0.12 | -1.78 | 0.076 | -0.44 | 0.02 |
|  | Time x Group: BD | 0.01 | 0.14 | 0.04 | 0.965 | -0.26 | 0.27 |
| Verbal memory | Intercept | 0.7 | 0.23 | 3.02 | **0.003** | 0.25 | 1.15 |
|  | Time | 0.15 | 0.08 | 1.9 | 0.058 | -0.01 | 0.31 |
|  | Group: SZ | -0.94 | 0.15 | -6.47 | **<0.001** | -1.23 | -0.66 |
|  | Group: BD | -0.48 | 0.16 | -3.01 | **0.003** | -0.8 | -0.17 |
|  | Sex: Male | -0.4 | 0.11 | -3.75 | **<0.001** | -0.61 | -0.19 |
|  | Age | -0.01 | 0.01 | -2.39 | 0.017 | -0.03 | 0.00 |
|  | Time x Group: SZ | -0.08 | 0.13 | -0.62 | 0.533 | -0.34 | 0.17 |
|  | Time x Group: BD | 0.17 | 0.15 | 1.17 | 0.241 | -0.12 | 0.47 |
| Semantic fluency | Intercept | -0.14 | 0.24 | -0.56 | 0.578 | -0.61 | 0.34 |
|  | Time | 0.06 | 0.07 | 0.92 | 0.358 | -0.07 | 0.20 |
|  | Group: SZ | -1.24 | 0.15 | -8.18 | **<0.001** | -1.53 | -0.94 |
|  | Group: BD | -0.36 | 0.16 | -2.21 | 0.027 | -0.68 | -0.04 |
|  | Sex: Male | -0.16 | 0.11 | -1.44 | 0.151 | -0.38 | 0.06 |
|  | Age | 0.01 | 0.01 | 1.01 | 0.315 | -0.01 | 0.02 |
|  | Time x Group: SZ | -0.28 | 0.12 | -2.37 | 0.018 | -0.51 | -0.05 |
|  | Time x Group: BD | 0.16 | 0.13 | 1.19 | 0.235 | -0.1 | 0.41 |
| Working memory | Intercept | 0.2 | 0.19 | 1.04 | 0.301 | -0.17 | 0.57 |
|  | Time | -0.11 | 0.06 | -1.67 | 0.095 | -0.23 | 0.02 |
|  | Group: SZ | -1.19 | 0.12 | -10.03 | **<0.001** | -1.42 | -0.96 |
|  | Group: BD | -0.8 | 0.13 | -6.12 | **<0.001** | -1.06 | -0.55 |
|  | Sex: Male | 0.14 | 0.09 | 1.57 | 0.118 | -0.03 | 0.31 |
|  | Age | -0.01 | 0.01 | -1.56 | 0.119 | -0.02 | 0.00 |
|  | Time x Group: SZ | 0.26 | 0.1 | 2.48 | 0.013 | 0.05 | 0.46 |
|  | Time x Group: BD | 0.35 | 0.12 | 2.93 | **0.003** | 0.12 | 0.59 |
| Cognitive control | Intercept | 0.26 | 0.24 | 1.09 | 0.277 | -0.2 | 0.72 |
|  | Time | 0.26 | 0.07 | 3.92 | **<0.001** | 0.13 | 0.38 |
|  | Group: SZ | -1.05 | 0.14 | -7.28 | **<0.001** | -1.33 | -0.77 |
|  | Group: BD | -0.62 | 0.16 | -3.94 | **<0.001** | -0.93 | -0.31 |
|  | Sex: Male | -0.34 | 0.11 | -3.13 | **0.002** | -0.55 | -0.13 |
|  | Age | 0.99 | 0.01 | -0.39 | 0.695 | -0.01 | 0.01 |
|  | Time x Group: SZ | 0.06 | 0.11 | 0.52 | 0.603 | -0.15 | 0.27 |
|  | Time x Group: BD | 0.19 | 0.12 | 1.52 | 0.129 | -0.05 | 0.42 |
| Abbreviations: SZ, schizophrenia; BD, bipolar disorder; HC, Health control; CRP, C-Reactive Protein; BMI, body mass index; SE, standardized error; CI, confidence interval [95%]. *Note*: Model estimates from linear mixed models for CRP and cognitive domains compares SZ and BD with HC. Estimates were considered significant if *p*<0.005 (Bonferroni). | | | | | | | |

| **Supplementary Table 7.** Subgroup comparisons of hierarchical clustering on SMI group alone, on sample and clinical characteristics at baseline | | | | |
| --- | --- | --- | --- | --- |
| **Characteristic** | **Cluster 1** N = 133^1^ | **Cluster 2** N = 75^1^ | **CI**^2^ | ***p***^3^ |
| Age | 26.7 (6.9) | 28.2 (8.8) | [-3.8, 0.9] | ns |
| Sex (female) | 74 (56%) | 26 (35%) | - | 0.004 |
| Education (years) | 13.4 (2.4) | 12.4 (2.3) | [0.2, 1.6] | 0.008 |
| WASI IQ (2-subtests) | 110.9 (12.1) | 98.8 (14.2) | [8.3, 16] | <0.001 |
| C-Reactive Protein | 0.9 (0.6) | 2.7 (1.9) | [-2.2, -1.3] | <0.001 |
| Composite, cognition | -0.3 (0.6) | -1.7 (0.9) | [1.1, 1.6] | <0.001 |
| BMI (kg/m²) | 23.9 (4.1) | 24.9 (4.4) | [-2.3, 0.2] | ns |
| PANSS Negative | 10.3 (5.0) | 12.5 (5.4) | [-3.7, -0.6] | 0.006 |
| PANSS Positive | 7.7 (3.9) | 9.1 (4.2) | [-2.5, -0.2] | 0.024 |
| PANSS Disorganized | 4.6 (2.0) | 5.5 (2.5) | [-1.6, -0.3] | 0.005 |
| PANSS Excited | 5.2 (1.6) | 5.1 (1.4) | [-0.2, 0.6] | ns |
| PANSS Depressed | 8.4 (3.0) | 8.4 (2.9) | [-0.8, 0.9] | ns |
| YMRS | 4.3 (4.6) | 4.9 (5.7) | [-2.2, 0.9] | ns |
| GAF Symptom | 53.2 (14.9) | 43.9 (12.9) | [5.4, 13] | <0.001 |
| GAF Function | 53.5 (14.4) | 43.1 (11.4) | [6.9, 14] | <0.001 |
| Duration of untreated illness | 102.9 (179.8) | 103.2 (220.3) | [-6, 63] | ns |
| Total, DDD | 1.6 (0.9) | 2.1 (1.2) | [-0.8, -0.1] | 0.006 |
| ^1^Mean (SD); n (%) ^2^CI = Confidence Interval, 95%  ^3^Welch Two Sample t-test; Pearson’s Chi-squared test  Abbreviations: WASI, Wechsler Abbreviated Scale of Intelligence; CRP, C-reactive Protein; BMI, body mass index; PANSS, Positive and Negative Syndrome Scale; YMRS, Young Mania Rating Scale; GAF, Global Assessment of Functioning scale; DDD, defined daily dosage. | | | | |
